# Forecast and interpretation of daily affected people during 21 days lockdown due to COVID 19 pandemic in India

**DOI:** 10.1101/2020.04.22.20075572

**Authors:** Achintya Bhattacharyya, Dipak Bhowmik, Joy Mukherjee

## Abstract

The problem of facing difficulty to control the spreading of newly detected novel corona virus 2019 is a matter of attention throughout the whole world. The total numbers of infected individuals have already crossed 2 million throughout the world. The government of all the affected countries have already taken many measures like lockdown to stop the spread. Therefore, it is important to study the nature of growth and interpretations are necessary for taking needful by the government. In this paper, we have tried to interpret the spreading capability of novel corona virus in India taking into consideration of 21 days lockdown data. The prediction is based on the present number of available cases and number of new cases reporting daily. We have explained the number of infected people in India comparing with Italy, China, Spain, and USA data of 21 days individually after announcing lockdown. The plot of daily new cases with the number of days spent after lockdown is fitted by linear and polynomial function. The best fitted graph is chosen for interpretation and that is based on the parameters obtained from fitting. The forecast of maximum spreading possibility is discussed with linear and 4-degree polynomial fitting parameters. Finally, some further preventive measurements are also discussed.

## Introduction

The COVID19 is a potentially severe acute respiratory infection caused by the corona virus 2(SARS-CoV-2). The physical manifestation consists of a respiratory infection with symptoms ranging from mild common cold to a severe viral pneumonia leading to acute respiratory distress that is potentially fatal. COVID-19 has been declared as a pandemic (panic-epidemic) on 11^th^ March by World Health Organization (WHO) all over the world [1]. The existence of corona virus is not new, but this time it has been spread from animal (bat) to human. The occurrence of having such epidemic had already been predicted last year in China [2]. The first spread of this novel corona virus was started from Wuhan, China on December, 2019. Now, almost all the parts of the world are exposed to this virus and since the inflectibility of this virus is large, a huge number of people have already become victim of it. In the lockdown situation, all the countries are trying to identify the infected people and isolate them. So, how many people will be more affected by this virus, is a million dollar question. For the countries like Italy, USA, Spain, India etc., the number of new cases are increasing day by day even after the declaration of lockdown by the respective government. The numbers are following a different trend of increasing for different country. Any natural phenomena are generally found to follow the exponential growth or decay, but in case of corona virus, the number of new cases per day is not at all exponential for all the countries. Many people have predicted the lockdown extension date by modeling the growth as exponential nature [3,4]. Hence, it is a challenging job to model the growth and to forecast the number of total affected people.

There has been a long history of research on epidemic deceases in statistical physics [5,6]. Simple mathematical models and fitting the variation with different polynomial can be useful for understanding and predicting the epidemic spreading. However, the accurate prediction of any epidemic spreading is very difficult due to the uncertainty of any modelling. So far some of the models show that the 49 days lockdown without any break could reach the baseline [7]. Also, sometimes the data may not be so reliable like the bird flu and SARS when the number of affected people and deaths were misrepresented to hide the extent of the epidemic. This type of problem may increase the uncertainty of the prediction and interpretation. Still, the scientific interpretation of the spreading of any epidemic deceases helps the government to understand its spreading and to take the necessary steps. The main problem of forecasting is that the general people fear about the forecasting and prediction of the spreading like Spanish flu which killed about 50 million people worldwide (1918-1920) and lasted almost 18 months. After that, no such big epidemic affected people like Spanish flu. COVID-19 is spreading the whole world like a way that it has affected more than 20 lacs people and killed more than 1.2 lacs people till now (15/04/2020). Despite this uncertainty, people have given model, prediction, and forecast on COVID 19 epidemic [8,9].

In this article, we use the data of worldometer [9] to interpret the growth in India with comparison of USA, China, Italy, and Spain. We have fitted the data of daily new cases over the days spent after lockdown in different functions like exponential, asymptotic and polynomial of different order. We have also compared the data with other countries like China, USA, Italy, Spain etc and tried to find out a trend of increasing or growing number of corona infected people mathematically. We observe that for China, the trend follows linear rather higher order polynomial and for USA, Italy, and Spain, it follows 4-degree polynomial fitting well. However, in case of India, it can be said that it has trend of both 4-degree polynomial and linear. Finally, we have predicted the underlying reason of maximum possible spreading of corona virus in India considering both linear and 4-degree polynomial fitting and suggested some preventive measures to stop this spreading.

## About COVID 19

The spreading of this virus has been declared pandemic as it is an infectious diseases that spreads quickly from person to person. The structure of the virus which has few possible qualities that allows spreading easily. Firstly, the virus is suggested to be a basically ball of proteins called spike proteins on the surface. Those spikes latch onto a protein called ACE 2 (Angiotensin-converting enzyme), which is found on the surface of human cells. The original SERS (Severe Acute Respiratory Syndrome) virus also did the same, but the latching onto ACE 2 is higher due to the specific shape of the spikes of SARS-CoV-2 because they are at closer fit to the ACE 2 protein. Once the attachment is made, the spike protein is split into two equal halves by an enzyme called Furin present in human body at large scale. Hence, the virus gets spread in human body. Most of the respiratory viruses tend to infect either the upper or lower airways of the respiratory system. If the upper respiratory system is infected by virus then the illness is very mild and if the lower airways is infected, they cause severe illness. SERS-Cov-2 seems to infect both the respiratory system due to the wide spread Furin enzyme. Thus, the virus causes severe contagiousness by attacking upper airways of respiratory system, which helps it to spread easily before moving to the lower one. According to the researcher including some research from Cambridge University, there are 3 types of COVID19 [11,12]. The variants are type A, B and C. The closest type of COVID 19 discovered in Bat was type A, the original human virus genome was presented in Wuhan. The mutated version of A was seen in American and was also found in the patients of USA. The A and C types are mainly found in Europeans and American country [12]. The B type is the most common type variant which is present in East Asia, and its ancestral genome appears not to have spread outside East Asia without mutation. In India, the type of this virus has not been publicly reported yet now. Being new with no vaccine and no adequate treatment protocol, it is challenging for the healthcare workers to combat.

## Analysis Method and discussions

In order to investigate the growth of the current epidemic COVID 19, we have taken consideration of 5 countries. China, being the source of the epidemic, made their lock down first on 23^rd^ of January. Following this Italy, Spain, USA, and India did the same on later time. Since the lockdown date is not same for all the countries, we have considered the first 21 days data after lockdown. We have collected the data from very authentic source “worldometer”. The new cases per day after lockdown announcement are plotted with the number of days since lockdown. Definitely, the five countries have different data but the variation of new cases per day with the no. of days seems to be almost identical fashion for all the countries except China. For better understanding of the nature, we have fitted the data for all the country linearly and 4-degree polynomial. We also tried to fit the data exponentially but that cannot be well fitted with that data point. The best fitted graphs are considered with those fitting covering the maximum data points. Figs. 1 (a)-(f) reveal the comparison of fitted data with one degree (linear) and 4 degree polynomial for Italy, China, and Spain. We see that in case of China, the data can be best fitted by a one-degree polynomial fit, i.e., linear fit, whereas the data points of other countries do not seem to have any signature of linearity. The variation of daily cases in Italy and Spain can be best fitted with 4 degree polynomial. Figs. 2 (a)-(d) show the same in case of USA and India. The data points of all the countries are best fitted using 4-degree polynomial. The coefficients of each degree are determined and used for further forecast of epidemicity of COVID 19.

**Fig. 1:**
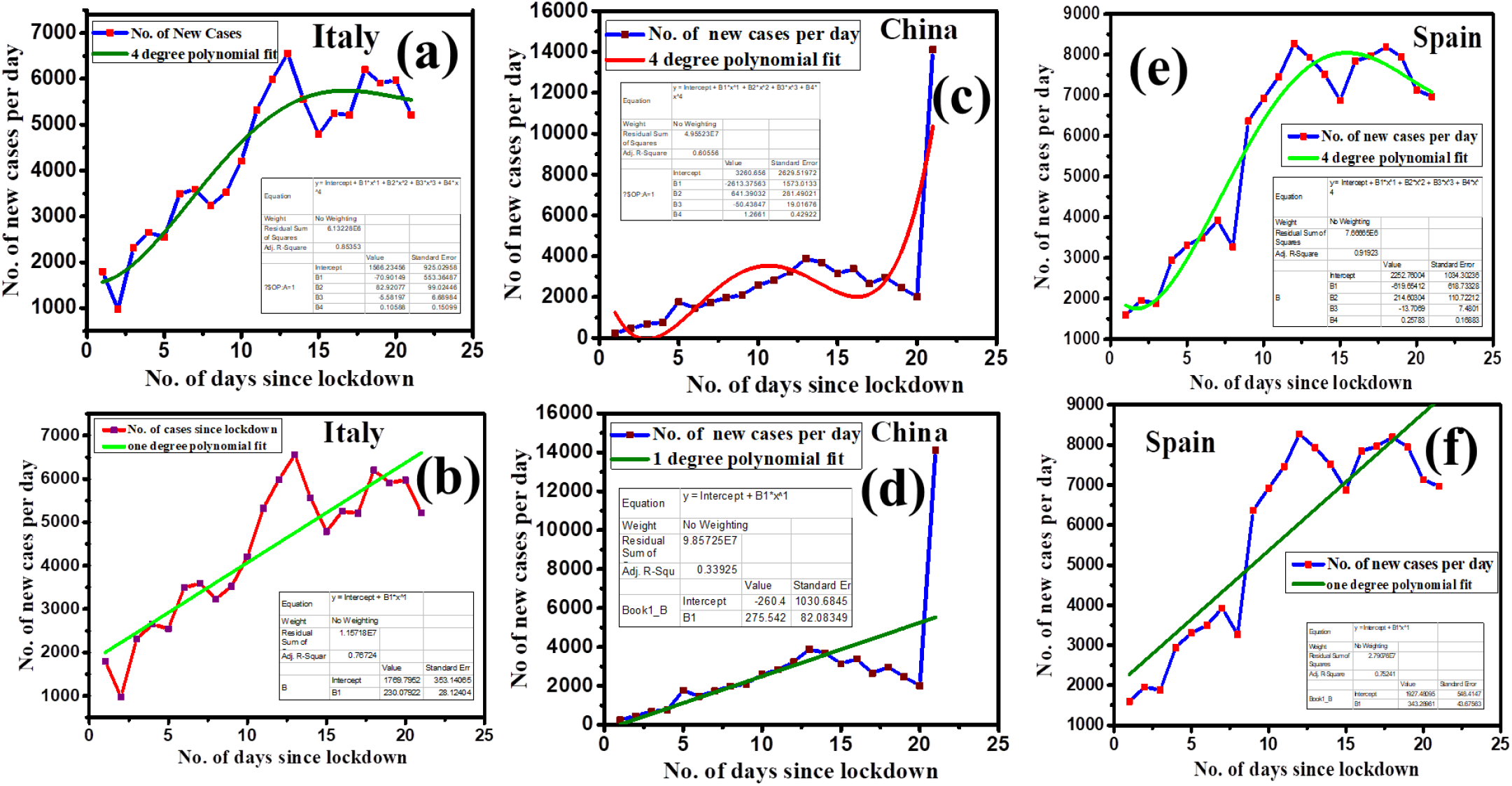
The 4-degree and one-degree polynomial fitting of the plot (no. of new cases per day vs. no of days since lockdown) for (a-b) Italy, (c-d) China, and (e-f) Spain, respectively. The fitting co-efficient and intercepts are shown in the legend of each plot.

**Fig. 2:**
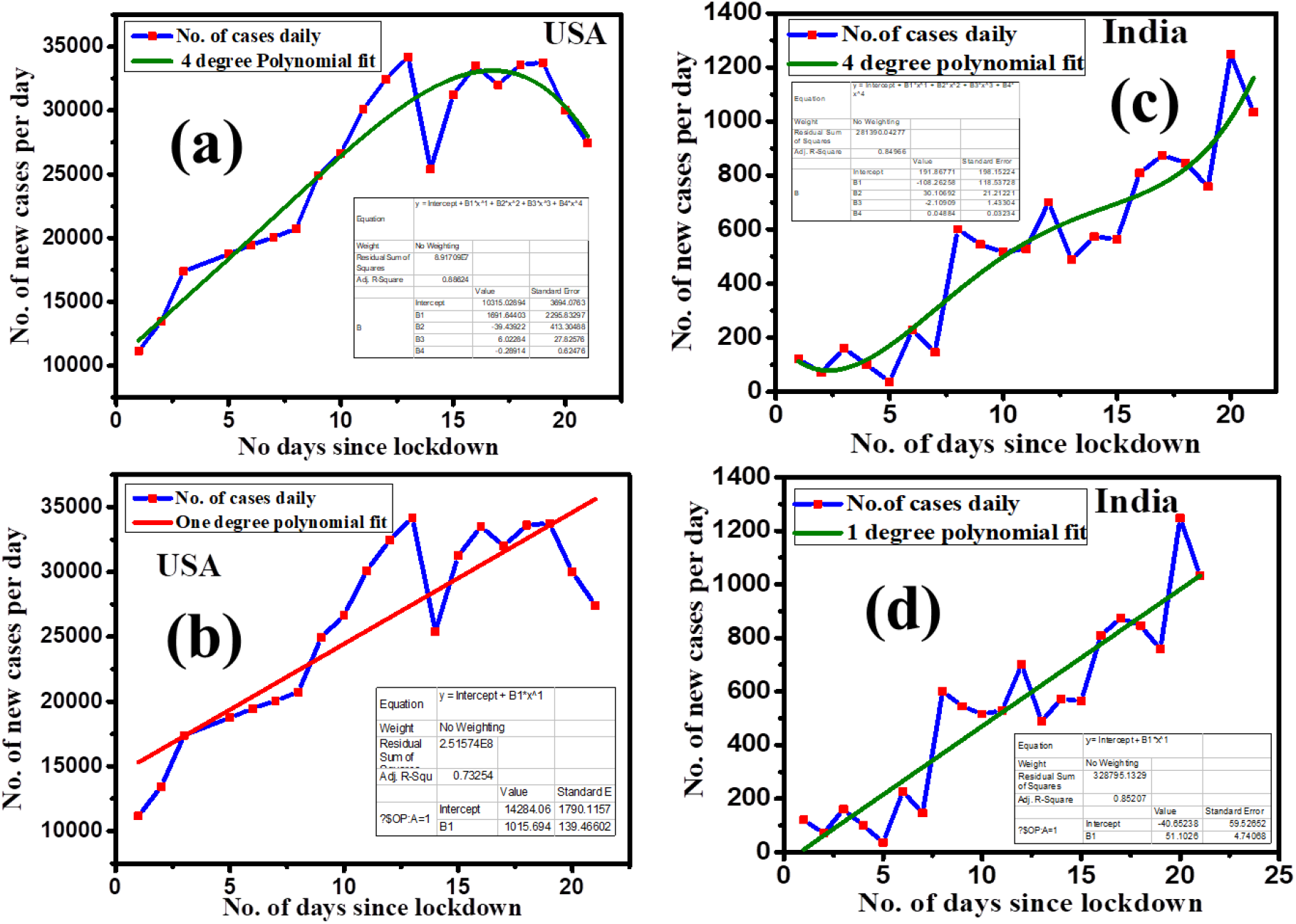
The 4-degree and one-degree polynomial fitting of the plot (no. of new cases per day vs. no of days since lockdown) for (a-b) USA and (c-d) India, respectively. The fitting co-efficient and intercepts are shown in the legend of each plot.

Before going into the details of fitting coefficient and their significance, let us discuss a natural process of spreading such corona virus. We know that the growth of any natural process follows exponential law. For example, if a person gets corona virus affected; whenever he/she comes into a country, he/she can spread *x* no. of people before going to quarantine. Now, the total number of affected people is (*1+x*). Again this *x* number of people will spread another *x* no. of people before going to quarantine. Hence, the total number will become (*1+x+x*^*2*^*+x*^*3*^*+x*^*4*^*+…)* = *e*^*x*^. Therefore, the number of affected people may grow exponentially for all the countries, but that is not the real case which we want to explore in this paper. In the case of China, the number of new cases grows almost linearly, whereas for the other country, it can be best fitted by a 4-degree polynomial. The coefficient of each degree of the polynomial will have different significance. Each of the country has their different coefficient and the number of people affected depends on these coefficients and the intercepts of y-axis (*Y*_*0*_), i.e., the number of initial affected people from lockdown. Let us consider a polynomial having degree 4, *Y(X) = Y*_*0*_*+B*_*1*_*X+B*_*2*_*X* ^*2*^*+B*_*3*_*X* ^*3*^*+B*_*4*_*X* ^*4*^. Here *x* is the days since lockdown and Y is the number of infected people reporting daily. The values of *Y*_*0*_, *B*_*1*,_ *B*_*2*,_ *B*_*3*,_ and *B*_*4*_ for different countries are listed below in Table 1. The terms *B*_*1*,_ *B*_*2*,_ *B*_*3*,_ and *B*_*4*_ correspond to the co-efficient of *X, X* ^*2*^, *X* ^*3*^ and *X* ^*4*^, respectively. The terms *X* ^*3*^ and *X* ^*4*^ are responsible for increasing daily +ve cases drastically indicating the community transmission, which are the crucial factor for spreading. So to control this epidemic, we must need to control the coefficient of different order of *X*. The main goal of preventing corona virus spreading is to attain minima of this graph with keeping the less number of days lockdown. To attain such minimum, the only way is to control the parameters *B*_*1*,_ *B*_*2*,_ *B*_*3*,_ and *B*_*4*_. If the coefficient of these two terms (*X* ^*3*^ and *X* ^*4*^) is less, the chance of community transmission for those countries is also less. These co-efficient are based on number of population, number of testing and how strict the lockdown are following. The variation of *X* ^*3*^ and *X* ^*4*^ are rapid and uncontrollable than that of *X* and *X* ^*2*^. Hence, the countries having lower value of this will reach to the baseline earlier than that having higher value.

**Table 1.** Values of the fitting co-efficient for different countries

| Country | $Y_0$ | $B_1$ | $B_2$ | $B_3$ | $B_4$ |
| --- | --- | --- | --- | --- | --- |
| Italy | 1566.23 | -70.90 | 82.92 | -5.58 | 0.10 |
| Spain | 2252.76 | -619.65 | 214.60 | -13.70 | 0.25 |
| USA | 10315.02 | 1691.64 | -39.43 | 6.02 | -0.29 |
| India | 191.86 | -108.26 | 30.10 | -2.10 | 0.049 |

Since, in case of China, the data cannot be fitted in a polynomial, hence the co-efficient values are not listed in Table 1. Here, it is very obvious that intersects (*Y*_*0*_) are different for different countries. This value mainly indicates the initial no. of infected people when the lockdown was declared. It has a significant role for predicting the spreading mechanism. If we look at the *Y*_*0*_ data from Table 1, we see that USA started lockdown after they reached the spread of infected people more than 10 thousands, then Spain, Italy, and India. For India, the value of *Y*_*0*_ is one order less compared to Italy, Spain, and USA. This was a very good decision by India government that India declared lockdown at early time when the number of infected people was not so large. For the higher number infected people when lockdown was declared in USA, the total number of infected people in USA is very high even after the lockdown period of 21 days than Spain and Italy. India has also one order less infected people till now even after 21 days lockdown period than Italy, Spain, and USA. We observe that for Italy, Spain, and India, the values of *B*_*2*_ and *B*_*4*_ are very important for predicting the number of affected people as *B*_*1*_ and *B*_*3*_ are negative for these countries. However, for USA, *B*_*1*_ and *B*_*3*_ are dominating factors. The values of *B*_*1*_ and *B*_*3*_ enhance the daily number of infected people and *B*_*2*_ and *B*_*4*_ decrease the number as these are negative for USA. In case of Italy, Spain, and India, *B*_*2*_ and *B*_*4*_ enhance the total number of people affected by corona virus. These co-efficient could well explain the number of affected people. In case of India, these co-efficient are less than that of Italy, Spain. So, the number of people infected in India is very less till now compared to Italy, Spain, and USA. However, we should not conclude that India will soon reach in baseline as the co-efficient of *X* ^*4*^ is still positive and it may increase. Also, in case of India, as the sufficient numbers of tests have not been conducted till now compared to Italy and Spain, the number of daily infected people may be increased certainly. If *B*_*4*_ increases due to withdrawing lockdown and not maintaining social distance in near future, then the community transmission effect may be very higher and it could be a dangerous situation than USA and other European country as the total population and its density in India are very high compared to other country. So, the lockdown may be withdrawn only after reducing sufficient number of daily +ve cases and social distancing should be maintained even after lockdown period for few months. The daily new cases in India can also be fitted linearly with the no of days since lockdown (Fig. 2d). It is good to visualize that the daily case in India can be saturated within a few days if the necessary steps are continued like China. The precautionary measures in medical background are discussed in next section.

## Preventive measures

As the forecast of spreading nature is not absolute, precautionary measures should always be taken until the spreading becomes almost at zero baseline. We have made some points as preventive measures which follow as

i. We should wash our hand with soap frequently. If the soap is not available, we should use alcohol based sanitizer.
ii. We should not touch eyes, mouth, and nose by our unclear hands as this virus can go inside our body through eyes, mouth, and nose.
iii. Every person should maintain at least 1 meter distance from any people.
iv. Social distancing (minimum 1 meter) should be maintained even after the lockdown period.
v. People should avoid gatherings, malls, gyms etc for certain periods.
vi. Special care should be taken during using our daily essential things like morning milk bags, elevators buttons, door bells, newspaper, car doors, raw vegetables and fruits, and shop counters.
vii. Random COVID 19 tests should be done in all localities even for asymptomatic people.
viii. Aerial Sanitization may be done and necessary vigilation should be done with drone for ensuring lockdown.
ix. The hotspot where the number of +ve cases is still increasing should be specially taken care.
x. The international as well as domestic travel by train, flight, bus may be kept suspended for some days even after lockdown period.

## Summary and conclusion

In summary, we have given the explanation of spreading the corona virus infection among the people by mathematical interpretation. The numbers of new cases are plotted with the number of days (21 days) after lockdown for India and some other countries for explaining the nature of the spreading. It is found that except China; Spain, Italy, USA follow the increase nature as 4-degree polynomial function. However, it is hopeful that India follows the increase nature as 4-degree polynomial with very low co-efficient values and it also follows the linear nature like China. Our analysis taking 21 days data after lockdown could explain the increase nature for USA, Spain, and Italy also. Finally, some preventive measures are discussed. We invite the researchers from all communities to come forward and work on COVID 19 according to their expertise. It may be helpful for fighting against this epidemic disease. Our analysis could give well explanation of daily affected people in last 21 days lockdown and also in near future. We strongly believe that India will recover soon from the present situation.

## Data Availability

All the data are available in the manuscript

## Acknowledgements

The authors (AB and JM) thank Variable Energy Cyclotron Centre Kolkata under Department of Atomic Energy, Govt. of India for providing financial support. One of the authors (DB) acknowledges Indian Institute of Technology Kanpur under Ministry of Human Resource Development (MHRD), Govt. of India for financial support. DB is grateful to Ms. Bhargabi Kundu for critical reading of the manuscript and fruitful suggestions. JM also thanks Ms. Rinku Mondol for valuable discussion.

## Authors Contribution

All authors contribute equally throughout the data analysis and paper writing.

## Data Availability Statement

All the data are available in the manuscript.

